# Mathematical Modeling and Simulation of SIR Model for COVID-2019 Epidemic Outbreak: A Case Study of India

**DOI:** 10.1101/2020.05.15.20103077

**Authors:** Ramjeet Singh Yadav

## Abstract

The present study discusses the spread of COVID-2019 epidemic of India and its end by using SIR model. Here we have discussed about the spread of COVID-2019 epidemic in great detail using Euler's method. The Euler’s method is a method for solving the ordinary differential equations. The SIR model has the combination of three ordinary differential equations. In this study, we have used the data of COVID-2019 Outbreak of India on 8 May, 2020. In this data, we have used 135710 susceptible cases, 54340 infectious cases and 1830 reward/removed cases for the initial level of experimental purpose. Data about a wide variety of infectious diseases has been analyzed with the help of SIR model. Therefore, this model has been already well tested for infectious diseases by various scientists and researchers. Using the data to the number of COVID-2019 outbreak cases in India the results obtained from the analysis and simulation of this proposed SIR model showing that the COVID-2019 epidemic cases increase for some time and there after this outbreak decrease. The results obtained from the SIR model also suggest that the Euler’s method can be used to predict transmission and prevent the COVID-2019 epidemic in India. Finally, from this study, we have found that the outbreak of COVID-2019 epidemic in India will be at its peak on 25 May 2020 and after that it will work slowly and on the verge of ending in the first or second week of August 2020.

## 1. Introduction

Today, the corona virus epidemic has emerged as an important challenge in front of the whole world. COVID-19 has about 3588777 confirmed cases and 247503 deaths as of May 6, 2020 [1]. Almost the entire population of the world is currently using lockdown, social distancing and masks to stop this epidemic. India is also using such resources to fight this epidemic at the moment. The COVID-19 epidemic is a member of SARS-Cov-2 family. No medicine has been prepared for this disease yet. COVD-2019 is an epidemic spreads from one human to another at a very rapid speed due to the breathing or contact of an infected person.

Hence COVID-2019 is a contagious disease. The incubation period of this disease is 2 to 14 days. In a recent study, it has been found that the overall mortality rate of COVID-19 epidemic is estimated at about 2–3%. This disease proves fatal for people above 60 years. The overall mortality rate for people above 40 years of age is about 27% [2, 3]. In India, on January 20, 2020, a patient of COVID-19 was found. This person came to Kerala of India from Wuhan city of China. The first case of COVID-19 was found at the end of November 2014 in Wuhan city of China. After 30 January 2020, the corona virus slowly spread in whole India.

On 19 March 2020, the Prime Minister of India, Narendra Modi announced the janata curfew on 22 March 2020. After this, the Prime Minister gave lockdown to India all over India till 14 April 2020. Even after lockdown in India, COVID-19 epidemic patients continued to grow. Today, according to COVID-19 epidemic data in India which was available on Indian council of medical research (ICMR) website, 81970 are infected, 27920 were cured and 2649 people died on 15 May 2020. However, India has a much larger population density than other countries and apart from this; medical facilities are not available in sufficient quantity. Therefore, the risk of spreading corona virus is very high here. Despite all these, corona infection in India is very less compared to other countries. In recent studies, it has been found that the cause of corona virus infection in India is low due to warm climate as well as humidity [4, 5], Bacille Calmette-Guérin (BCG) vaccination and a large amount of young population [6]. Due to all this reasons, the resistance of people here is very high compared to other countries. All these studies are preliminary studies and no scientific evidence of this type of study is available till now [7]. Hence there is a need to study COVID-2019 outbreak with more evidence now. In this study we have presented an epidemic model based on SIR method of COVID-19 spread to India. Most epidemics have an initial exponential curve and then gradually flatten out [8]. In this proposed study, we have also considered the effects of social distancing on the growth of infections, lockdown and face mask India. India announced a countrywide lockdown on 24 March 2020 for 21 days although a study has suggested that this period may be insufficient for controlling the COVID-19 pandemic [9]. In the present study, we have assumed the effects of social distancing measures, lockdown and face cover from the time of spread to India.

Hence there is a need to study COVID-2019 with more evidence now. In this proposed study, we have presented an epidemic model based on SIR method of COVID-19 spread to India. The proposed SIR model has three differential equations. The solution of such type of differential equation is difficult and time consuming. Therefore we have used Euler’s method for solving these three differential equations. Most epidemics have an initial exponential curve and then gradually flatten out.

The objectives of these studies are given below:

1. Finding the rate of spread of the disease with help of SIR model.
2. The development SIR model for exposed COVID-2019 outbreak at peak in India.
3. Forecast of COVID-2019 outbreak of India with next days, months even a year for better management for doctors and various government organizations.
4. For find out the ending stage of COVID-2019 outbreak in India.

## 2. SIR Model

In this proposed study, we have considered an epidemic model which was developed by Kermack and McKendrick in 1927 [10]. This epidemic model is also known as SIR (Susceptible, Infective and Recover/Removed) epidemic model. This model have already used successfully in several outbreak diseases like Avian influenza, Cholera, SARS, Ebola, Plague, Yellow fever, Meningitis, MERS, Influenza, Zika, Rift Valley Fever, Lassa fever, Leptospirosis [11, 12, 13, 14, 15]. The SIR model is very useful for future prediction, end and peak of epidemic disease and other related activity of outbreak diseases [12].

Let us consider the population of India remains constant regarding the study COVID-2019 outbreak in India. Here, we have chosen all COVID-2019 tested population of India on 7 April 2020. In this proposed study, we have total COVID-2020 tested population is divided into three parts:

1. *S*(*t*): The number of susceptible population at time *t* i.e. number of total COVID-2019 tested population till 7 April 2020.
2. *I*(*t*): The number of infectives population at time *t*, i.e. number of infected COVID-2019 population of India till 7 April 2020.
3. *R*(*t*): The number of recovered population at time *t*, i.e. number of recovered or died or naturally immune to the disease COVID-2019 population of India till 7 April 2020.

In this proposed study, we have take R(t) is equal to the recovered population plus died population from COVID-2019 outbreak of India on 7 April 2020 for the sake of simplicity of this study [16]. Figure 1 shows the description of proposed SIR model for not considering virus evolution.

**Figure 1:**
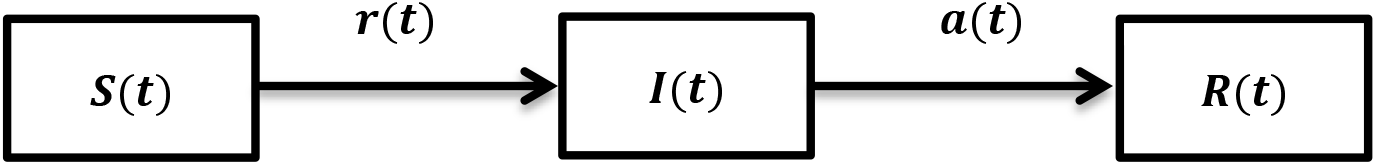
Description of SIR model not considering COVID-2019 outbreak virus evolution

This model does not consider the development of COVID-2019 like most of the diseases. But, in contrast my proposed SIR model which is shown in figure 2 does consider the development of COVID-2019 outbreak of India. This model also predicts maximum growth of COVID-2019 outbreak in India. Figure 2 shows the description of SIR model for recovered re-tuning into susceptible because the COVID-2019 outbreak of India has evolved into one which can re-infect.

**Figure 2:**
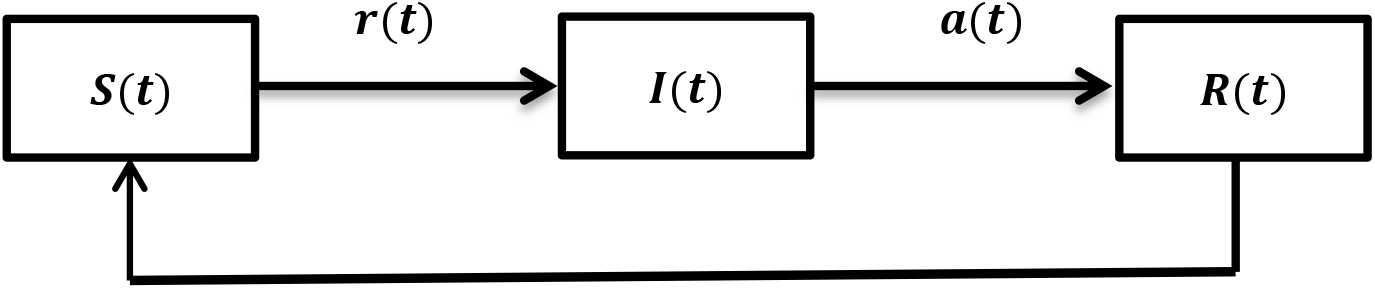
Description of SIR model considering COVID-2019 outbreak virus evolution

## 3. Methodology of SIR Model

Let us consider the following three differential equations are used for experimental studies and experimental discussion for COVID-2019 of India. The description of these three differential equations is given below:

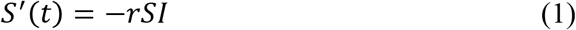

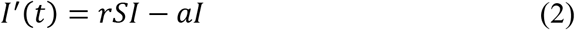

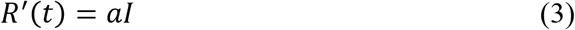

The parameters *r* and *a of* above differential equations are known as the infection rate and recovery/removal rate of COVID-2019 of India. In this proposed study the average time of COVID-2019 outbreak of India is approximately 14 days. These numerical values of *r* and *a* are very useful in initial level for solving the three differential equations of COVID-2019 outbreak of India.

The three differential equations (1), (2) and (3) of the proposed SIR epidemic model for COVID-2020 outbreak of India can be also written as [12]:

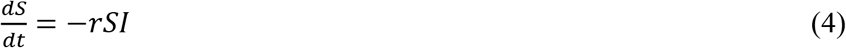

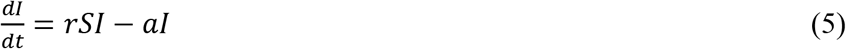

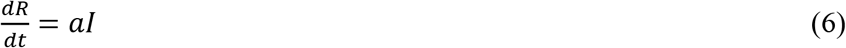

These three differential equations of SIR model is known as Kermack-McKendrick [12] SIR model. At the present time, this model is very useful for the data analysis of COVID-2019 in India. Again adding equation number (4), (5) and (6), we can get another very useful expression for COVID-2019 data analysis. This expression is given below:

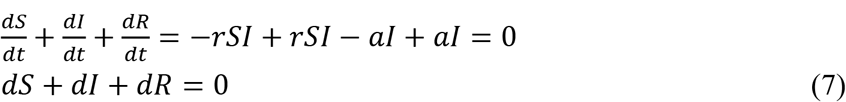

After integrating equation number (7), we can get the following relation for calculating the total population of COVID-2019:

*S*′ + *I*′ + *R*′ = *N*, where *N* is known as the constant of integration which is measure the total size of population for COVID-2019 at initial level and after end the epidemic COVID-2019 in India. This is constant population at all levels of COVID-2019 outbreak. The above expression *S*′ + *I*′ + *R*′ = *N* can be also denoted by in the following form:

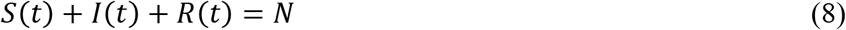

For the experimental purpose of data analysis of COVID-2019 outbreak of India, we can take the following initial values of proposed SIR model, i.e.

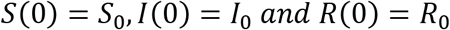

Here the population size of India is constant. We can calculate the recovered population of COVID-2019 outbreak of India which given by the following formula:

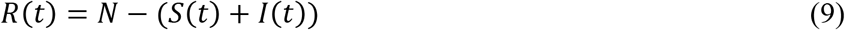

The above three differential equations (4), (5) and (5) of the proposed SIR model can be converted into two differential equations equation number (9). The solution of these two differential equations is very difficult and time consuming. But the solution is very necessary of these two differential equations for data analysis of COOVID-2019 outbreak of India. In this proposed study, we have used quantitative approach for solving these two differential equations of SIR model.

Now, here we can say that if *S*′ is less than zero for all *t* and if *I*′ is greater than zero as long as the initial population (say the number of susceptible cases in India on 7 May 2020) *S*_0_ is greater than the ratio, 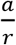. In other words, we can say that we will initially increase to some r maximum if initial population *S*_0_ is greater than the ratio 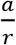, but eventually it must decrease r and approaching to zero because *S*_0_ decreasing. In this proposed study, we have introduced some cases for COVID-2019 outbreak of India, which is given below:

### Case-1

If *S*_0_ is less than the ratio, 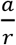 then the infection *I* of COVID-2020 outbreak of India r will be decrease or simply to be zero after some times.

### Case-2

If *S*_0_ is greater than the ratio 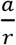 then the infection *I* of COVID-2020 outbreak of India r will be epidemic of COVID-2019.

These are the assumptions of SIR model regarding the COVID-2019 outbreak of India. Therefore from the above two assumptions, we can say that the behavior of COVID-2019 outbreak of India depends on the values of following expression: n

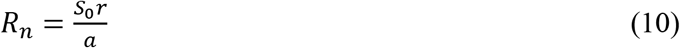

This quantity is known as the threshold number. In this present study we have defined another quantity called reproductive number which is denoted by *R*_n_ and defined by the following expression (10). This is the number of secondary infectives of COVID-2019 outbreak produced by one primary infective in the susceptible populations. Here, there are two cases of COVID-2019 of India regarding reproductive number:

#### Case-1

If R_n_ is less than one then COVID-2019 outbreak will be does out from India.

#### Case-2

If R_n_ is more than one, then the outbreak of COVID-2019 is still in epidemic form in India.

### 3.1. Phase Plan and Experimental Results of COVID-2019 Outbreak of India

There is an absolute need to solve the differential equation of the proposed the SIR model for analysis of COVID-2019 outbreak of India. Let us consider a population of susceptible of COVID-2019 outbreak and a small number of infected populations. Is the of COVID-2019 infectives populations increase substantially in India? The answer of this question will get after solving differential equations of (4), (5) and (6). The differential equations (4), (5) and (6) is system of differential equation and these equations have three unknown. These systems of differential equations are very difficult to solve. Although, after combining the equation (4) and (5) then we get the single differential equation with one unknown for the proposed SIR model. The procedure is as follows:

According to the chain rule calculus:

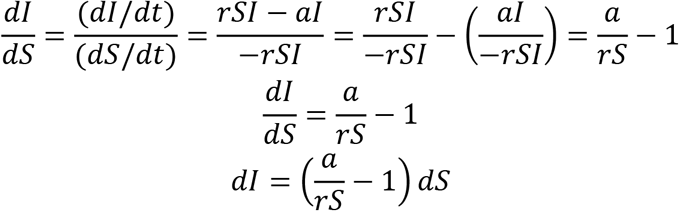

Integrating both sides of above equation, we get

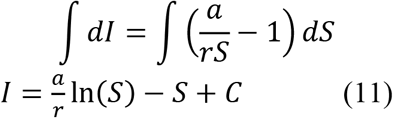

Where, C is the arbitrary constant.

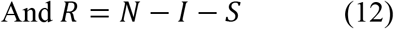

This Karmack-Mchendrick SIR model is equipped with the initial conditions. We take the initial conditions which are given below:

*S*(0) = *S*_0_ and *I*(0) = I_0_ then the equation (11) becomes:

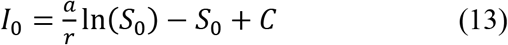

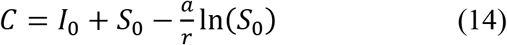

Let us consider the population size of susceptible case of COVID-2019 outbreak of India is *K*. This is approximately equal to initial population S_0_ of India. Here, we will introduce a small number of infectives in the population. Therefore,

*S*_0_ = K, I_0_ = 0 and 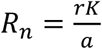

If *I*(*t*) = 0 as *t* → ∞ and 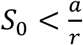 then *V*(*S*_0_, *I*_0_) = *V*(*S*_0_), gives the following expression:

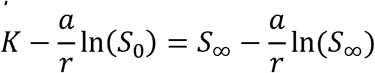

Where *S*_∞_ is the susceptible population of India if infectives case will be zero. After simplification of above expression, we will get the following expression:

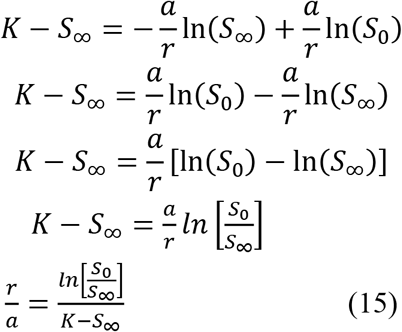

Here 0 < *S*_∞_ < *K* that is past of the population of India escapes the COVID-2019 infective. In this proposed study, it is very difficult to estimate the parameters of *r* and *a* because these are depends on disease being studies and on social and behavioral factors of that country. The population *S*_∞_ and *S*_0_ can be estimated by serological studies before and after of the COVID-2019 outbreak and using this data, the basic reproduction number is given by the following formula:

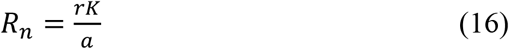

This expression can be calculated using expression (15). The maximum number of COVID-2019 outbreak infectives at any time in India can be obtained by substantially using the following calculation:

Putting 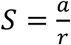 and *I* = *I*_max_ in equation (11), we get the maximum number of infective case of COVID-2019 outbreak in India at any time.

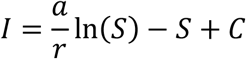

Where, 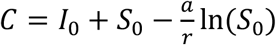

Therefore the maximum number of infectives cases *I*_*max*_ of COVID-2019 outbreak of India can calculated with the help of following expression:

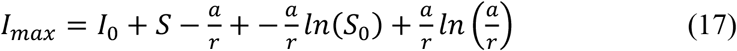

The differential equation of the proposed SIR model can be solved with help of many numerical methods such as Runge Kutta and Euler methods. Here we have used Euler method for solving SIR model based differential equation.

In this proposed study, we have used the MATLAB software for solving the differential equation using the above initial conditions values of *S*_0_, *I*_0_, *R*_0_, *a* and *r*. The experimental results of SIR model is shown in table 1. Here, the numerical calculation and data analysis of COVID-2019 outbreak of India has been done with the help of Euler method.

Euler’s method is purely numerical method for solving the first order differential equations. The SIR model has also system of first order differential equations. So, the Euler’s method is more suitable for solving the proposed SIR based system of differential equations. The description of the Euler’s method is given below:

Let consider the first order differential equation:

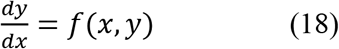

The solution of differential equation (18) is given by the following expression:

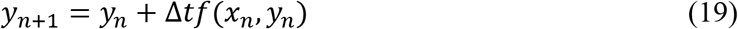

Where ∆*t* is a small step size in the time domain and *f*(*x*_n_, *y*_n_) is the slope of the curve. Here, we want to calculate the dependent variable called S, I and R to the proposed SIR model. Therefore the solution of proposed SIR model based differential is converted into Euler method forms which are given below:

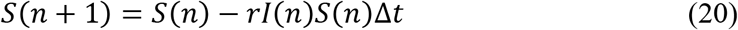

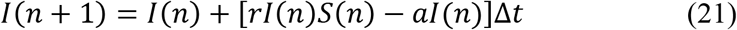

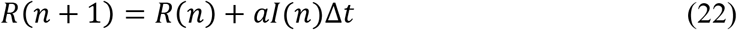

In this proposed study, we have used COVID-2019 data set from India on 7 May 2020. Here, we have taken the total number of COVID-2019 tested population as *S*_0_, total number of infectives population as *I*_0_ and total number of recovered/removed cases as R_0_ at initial level for analyzing the COVID-2019 outbreak of India on 7 May 2020. These three initial populations *S*_0_, *I*_0_ and *R*_0_ are represented as:

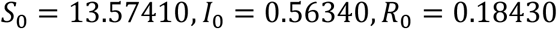

The value of recovery rate/removal rate and infection rate of COVID-2019 outbreak of India can be calculated with the help by the following expression:

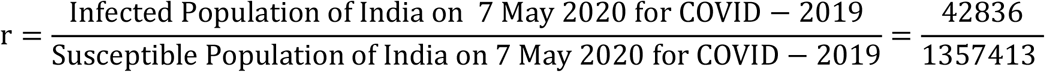

*r* = 0.03156

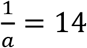, (Because the incubation time of COVID-2019 outbreak of India is 14 day)

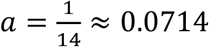

∆*t* = 0.1407

Putting the values of ∆*t*, *r, a*, *S*_0_, *I*_0_ and R_0_ in equation (20), (21) and (22) to get the next generation values Susceptible population *S*_1_, *I*_1_ and *R*_1_,

*S*_1_ = *S*_0_ − *rI*_0_*S*_0_∆*t*

*S*_1_ = 13.57410 − (0.03156 × 0.56340 × 13.57410 × 0.1407)

*S*_1_ = 13.57410 − 0.03396

*S*_1_ = 13.54014

*I*_1_ = *I*_0_ + (*rI*_0_*S*_0_ − *aI*_0_)∆*t*

*S*_1_ = 0.56340 + (0.03156 × 0.56340 × 13.57410 × 13.57410 − 0.07143 × 0.56340) × 0.1407

*S*_1_ = 0.591698

*R*_1_ = *R*_0_ + *aI*_0_∆*t*

*R*_1_ = 0.18430 + 0.07143 × 0.56340 × 0.1407

*R*_1_ = 0.1899622832434

*R*_1_ = 0.18996

Similarly, we can calculate other iteration. The numerical results of Euler’s method of SIR model is shown table 1.

**Table 1:** SIR Methods Simulation and Results of Runge Kutta Fourth Order Method.

| S.No. | Date | Day/Time | S | I | R | S+I+R |
| --- | --- | --- | --- | --- | --- | --- |
| 1 | 7-May-2020 | 0 | 13.57410 | 0.56340 | 0.18430 | 14.32180 |
| 2 | 8-May-2020 | 0.1407 | 13.53930 | 0.59240 | 0.19010 | 14.32180 |
| 3 | 9-May-2020 | 0.2815 | 13.50290 | 0.62280 | 0.19620 | 14.32190 |
| 4 | 10-May-2020 | 0.4222 | 13.46460 | 0.65460 | 0.20260 | 14.32180 |
| 5 | 11-May-2020 | 0.5629 | 13.42450 | 0.68790 | 0.20930 | 14.32170 |
| 6 | 12-May-2020 | 1.2091 | 13.21490 | 0.86190 | 0.24500 | 14.32180 |
| 7 | 13-May-2020 | 1.8552 | 12.95740 | 1.07490 | 0.28950 | 14.32180 |
| 8 | 14-May-2020 | 2.5013 | 12.64410 | 1.33280 | 0.34490 | 14.32180 |
| 9 | 15-May-2020 | 3.1474 | 12.26760 | 1.64090 | 0.41330 | 14.32180 |
| 10 | 16-May-2020 | 3.99 | 11.67150 | 2.12430 | 0.52600 | 14.32180 |
| 11 | 17-May-2020 | 4.8326 | 10.94780 | 2.70320 | 0.67090 | 14.32190 |
| 12 | 18-May-2020 | 5.6752 | 10.09940 | 3.36910 | 0.85330 | 14.32180 |
| 13 | 19-May-2020 | 6.5178 | 9.14520 | 4.09880 | 1.07780 | 14.32180 |
| 14 | 20-May-2020 | 7.5519 | 7.87820 | 5.02850 | 1.41510 | 14.32180 |
| 15 | 21-May-2020 | 8.586 | 6.58980 | 5.91230 | 1.81970 | 14.32180 |
| 16 | 22-May-2020 | 9.6202 | 5.36510 | 6.67150 | 2.28520 | 14.32180 |
| 17 | 23-May-2020 | 10.6543 | 4.27120 | 7.24990 | 2.80070 | 14.32180 |
| 18 | 24-May-2020 | 11.8923 | 3.19080 | 7.66910 | 3.46190 | 14.32180 |
| **19** | **25-May-2020** | **13.1304** | **2.35370** | **7.82000** | **4.14810** | **14.32180** |
| 20 | 26-May-2020 | 14.3684 | 1.72990 | 7.75340 | 4.83860 | 14.32190 |
| 21 | 27-May-2020 | 15.6065 | 1.28090 | 7.52520 | 5.51570 | 14.32180 |
| 22 | 28-May-2020 | 16.5352 | 1.03080 | 7.28400 | 6.00700 | 14.32180 |
| 23 | 29-May-2020 | 17.4639 | 0.83580 | 7.00500 | 6.48100 | 14.32180 |
| 24 | 30-May-2020 | 18.3926 | 0.68320 | 6.70270 | 6.93590 | 14.32180 |
| 25 | 31-May-2020 | 19.3213 | 0.56360 | 6.38800 | 7.37030 | 14.32190 |
| 26 | 1-June-2020 | 20.2499 | 0.46950 | 6.06890 | 7.78340 | 14.32180 |
| 27 | 2-June-2020 | 21.1786 | 0.39480 | 5.75160 | 8.17540 | 14.32180 |
| 28 | 3-June-2020 | 22.1073 | 0.33510 | 5.44010 | 8.54660 | 14.32180 |
| 29 | 4-June-2020 | 23.036 | 0.28690 | 5.13750 | 8.89740 | 14.32180 |
| 30 | 5-June-2020 | 24.1739 | 0.24010 | 4.78130 | 9.30040 | 14.32180 |
| 31 | 6-June-2020 | 25.3118 | 0.20350 | 4.44320 | 9.67510 | 14.32180 |
| 32 | 7-June-2020 | 26.4497 | 0.17450 | 4.12410 | 10.02320 | 14.32180 |
| 33 | 8-June-2020 | 27.5876 | 0.15130 | 3.82440 | 10.34610 | 14.32180 |
| 34 | 9-June-2020 | 29.0546 | 0.12780 | 3.46620 | 10.72780 | 14.32180 |
| 35 | 10-June-2020 | 30.5215 | 0.10970 | 3.13850 | 11.07360 | 14.32180 |
| 36 | 11-June-2020 | 31.9885 | 0.09550 | 2.83970 | 11.38660 | 14.32180 |
| 37 | 12-June-2020 | 33.4554 | 0.08430 | 2.56780 | 11.66970 | 14.32180 |
| 38 | 13-June-2020 | 35.604 | 0.07170 | 2.21410 | 12.03600 | 14.32180 |
| 39 | 14-June-2020 | 37.7526 | 0.06240 | 1.90760 | 12.35180 | 14.32180 |
| 40 | 15-June-2020 | 39.9012 | 0.05540 | 1.64270 | 12.62370 | 14.32180 |
| 41 | 16-June-2020 | 42.0498 | 0.05000 | 1.41410 | 12.85780 | 14.32190 |
| 42 | 17-June-2020 | 45.092 | 0.04420 | 1.14270 | 13.13500 | 14.32190 |
| 43 | 18-June-2020 | 48.1343 | 0.04000 | 0.92290 | 13.35890 | 14.32180 |
| 44 | 19-June-2020 | 51.1766 | 0.03700 | 0.74560 | 13.53910 | 14.32170 |
| 45 | 20-June-2020 | 54.2188 | 0.03480 | 0.60240 | 13.68470 | 14.32190 |
| 46 | 21-June-2020 | 57.0367 | 0.03310 | 0.49390 | 13.79480 | 14.32180 |
| 47 | 22-June-2020 | 59.8547 | 0.03180 | 0.40500 | 13.88500 | 14.32180 |
| 48 | 23-June-2020 | 62.6726 | 0.03080 | 0.33210 | 13.95890 | 14.32180 |
| 49 | 24-June-2020 | 65.4905 | 0.03000 | 0.27240 | 14.01940 | 14.32180 |
| 50 | 25-June-2020 | 68.2881 | 0.02940 | 0.22360 | 14.06880 | 14.32180 |
| 51 | 26-June-2020 | 71.0857 | 0.02880 | 0.18350 | 14.10940 | 14.32170 |
| 52 | 27-June-2020 | 73.8833 | 0.02840 | 0.15070 | 14.14270 | 14.32180 |
| 53 | 28-June-2020 | 76.6809 | 0.02810 | 0.12380 | 14.17000 | 14.32190 |
| 54 | 29-June-2020 | 79.4708 | 0.02780 | 0.10160 | 14.19240 | 14.32180 |
| 55 | 30-June-2020 | 82.2608 | 0.02760 | 0.08350 | 14.21080 | 14.32190 |
| 56 | 1-July-2020 | 85.0508 | 0.02740 | 0.06860 | 14.22590 | 14.32190 |
| 57 | 2-July-2020 | 87.8408 | 0.02720 | 0.05630 | 14.23820 | 14.32170 |
| 58 | 3-July-2020 | 90.6279 | 0.02710 | 0.04630 | 14.24840 | 14.32180 |
| 59 | 4-July-2020 | 93.415 | 0.02700 | 0.03800 | 14.25680 | 14.32180 |
| 60 | 5-July-2020 | 96.2022 | 0.02690 | 0.03120 | 14.26360 | 14.32170 |
| 61 | 6-July-2020 | 98.9893 | 0.02690 | 0.02570 | 14.26930 | 14.32190 |
| 62 | 7-July-2020 | 101.775 | 0.02680 | 0.02110 | 14.27390 | 14.32180 |
| 63 | 8-July-2020 | 104.561 | 0.02680 | 0.01730 | 14.27770 | 14.32180 |
| 64 | 9-July-2020 | 107.347 | 0.02670 | 0.01420 | 14.28090 | 14.32180 |
| 65 | 10-July-2020 | 110.133 | 0.02670 | 0.01170 | 14.28340 | 14.32180 |
| 66 | 11-July-2020 | 112.918 | 0.02670 | 0.00960 | 14.28550 | 14.32180 |
| 67 | 12-July-2020 | 115.704 | 0.02670 | 0.00790 | 14.28730 | 14.32190 |
| 68 | 13-July-2020 | 118.489 | 0.02660 | 0.00650 | 14.28870 | 14.32180 |
| 69 | 14-July-2020 | 121.275 | 0.02660 | 0.00530 | 14.28990 | 14.32180 |
| 70 | 15-July-2020 | 124.06 | 0.02660 | 0.00440 | 14.29080 | 14.32180 |
| 71 | 16-July-2020 | 126.845 | 0.02660 | 0.00360 | 14.29160 | 14.32180 |
| 72 | 17-July-2020 | 129.63 | 0.02660 | 0.00300 | 14.29230 | 14.32190 |
| 73 | 18-July-2020 | 132.415 | 0.02660 | 0.00240 | 14.29280 | 14.32180 |
| 74 | 19-July-2020 | 135.2 | 0.02660 | 0.00200 | 14.29320 | 14.32180 |
| 75 | 20-July-2020 | 137.985 | 0.02660 | 0.00160 | 14.29360 | 14.32180 |
| 76 | 21-July-2020 | 140.77 | 0.02660 | 0.00130 | 14.29390 | 14.32180 |
| 77 | 22-July-2020 | 143.555 | 0.02660 | 0.00110 | 14.29410 | 14.32180 |
| 78 | 23-July-2020 | 146.34 | 0.02660 | 0.00090 | 14.29430 | 14.32180 |
| 79 | 24-July-2020 | 149.125 | 0.02660 | 0.00070 | 14.29450 | 14.32180 |
| 80 | 25-July-2020 | 151.91 | 0.02660 | 0.00060 | 14.29460 | 14.32180 |
| 81 | 26-July-2020 | 154.695 | 0.02660 | 0.00050 | 14.29470 | 14.32180 |
| 82 | 27-July-2020 | 157.48 | 0.02660 | 0.00040 | 14.29480 | 14.32180 |
| 83 | 28-July-2020 | 160.265 | 0.02660 | 0.00030 | 14.29490 | 14.32180 |
| 84 | 29-July-2020 | 163.05 | 0.02660 | 0.00030 | 14.29500 | 14.32190 |
| 85 | 30-July-2020 | 165.835 | 0.02660 | 0.00020 | 14.29500 | 14.32180 |
| 86 | 31-July-2020 | 169.03 | 0.02660 | 0.00020 | 14.29510 | 14.32190 |
| 87 | 1-August-2020 | 172.225 | 0.02660 | 0.00010 | 14.29510 | 14.32180 |
| 88 | 2-August-2020 | 175.42 | 0.02660 | 0.00010 | 14.29510 | 14.32180 |
| 89 | 3-August-2020 | 178.615 | 0.02660 | 0.00010 | 14.29510 | 14.32180 |
| 90 | 4-August-2020 | 182.323 | 0.02660 | 0.00010 | 14.29520 | 14.32190 |
| **91** | **5-August-2020** | **186.031** | **0.02660** | **0.00010** | **14.29520** | **14.32190** |
| 92 | 6-August-2020 | 189.739 | 0.02660 | 0.00000 | 14.29520 | 14.32180 |
| 93 | 7-August-2020 | 193.448 | 0.02660 | 0.00000 | 14.29520 | 14.32180 |
| 94 | 8-August-2020 | 195.086 | 0.02660 | 0.00000 | 14.29520 | 14.32180 |
| 95 | 9-August-2020 | 196.724 | 0.02660 | 0.00000 | 14.29520 | 14.32180 |
| 96 | 10-August-2020 | 198.362 | 0.02660 | 0.00000 | 14.29520 | 14.32180 |
| 97 | 11-August-2020 | 200.000 | 0.02660 | 0.00000 | 14.29520 | 14.32180 |

Figure 3 shows simulation of proposed SIR Model for COVID-2019 epidemic state of India from 7-May 2020. This figure also shows that the date of the maximum number of infection cases of COVID-2019 in India is 25 May 2020 (see table 1 from bold column). The figure 4 shows the maximum number of infected cases of COVID-2019 outbreak of India. Apart from this, the figure 5 shows recovered cases of COVID-2019 outbreak of India.

**Figure 3:**
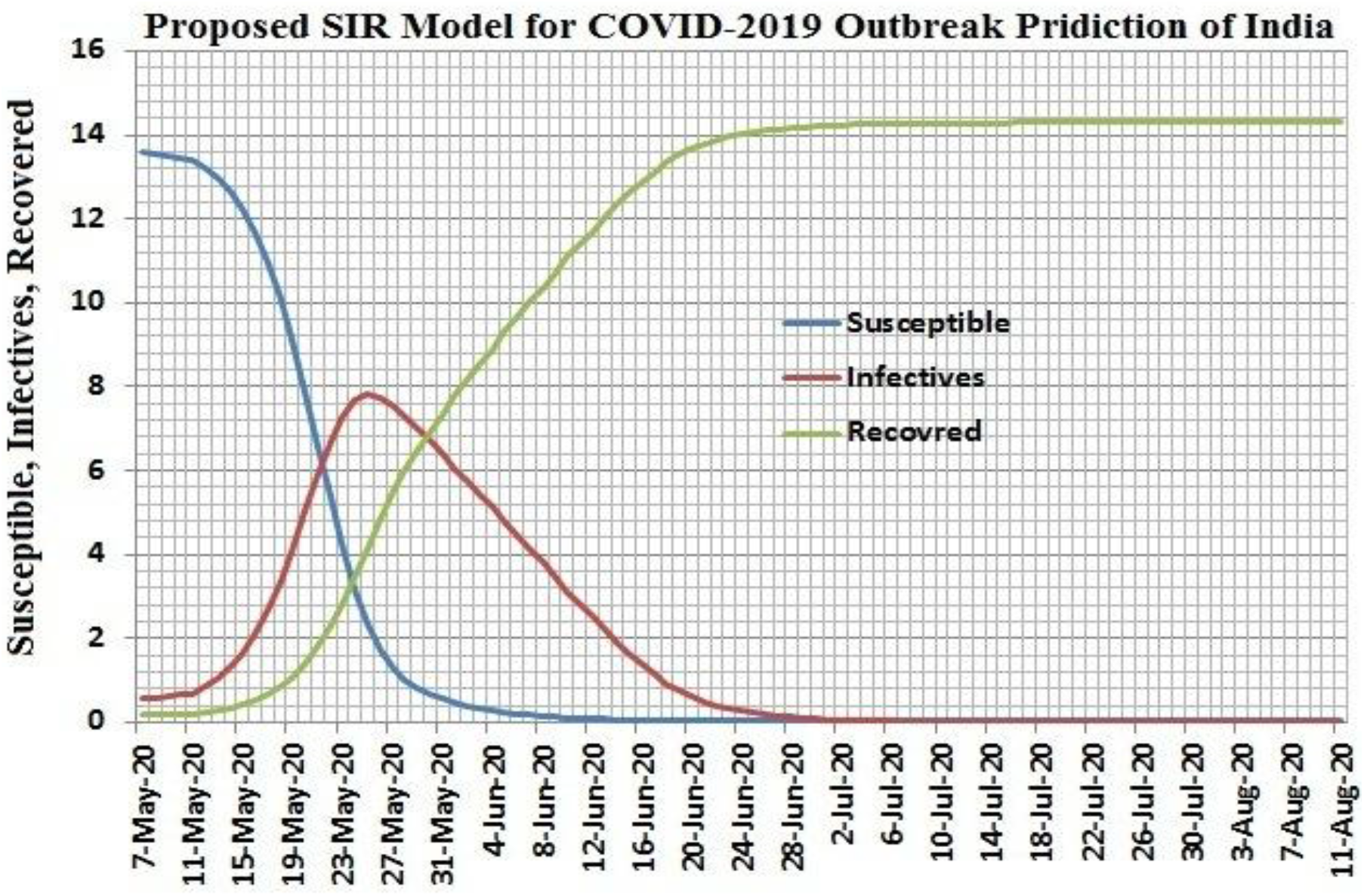
SIR Model Simulation for COVID-2019 epidemic state of India from 7-May 2020

**Figure 4:**
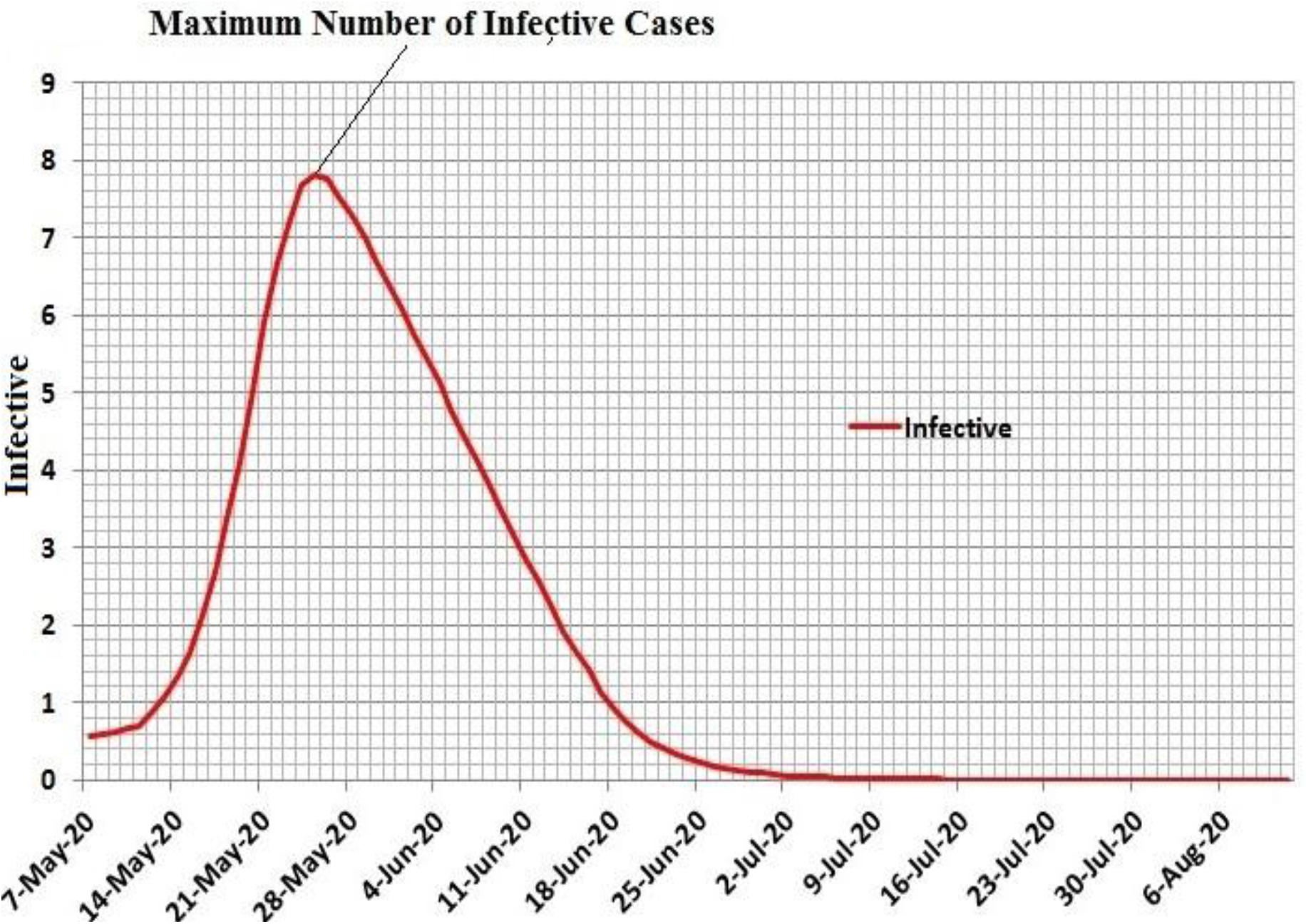
Maximum Number of Infective cases of COVI-2019 outbreak of India

**Figure 5:**
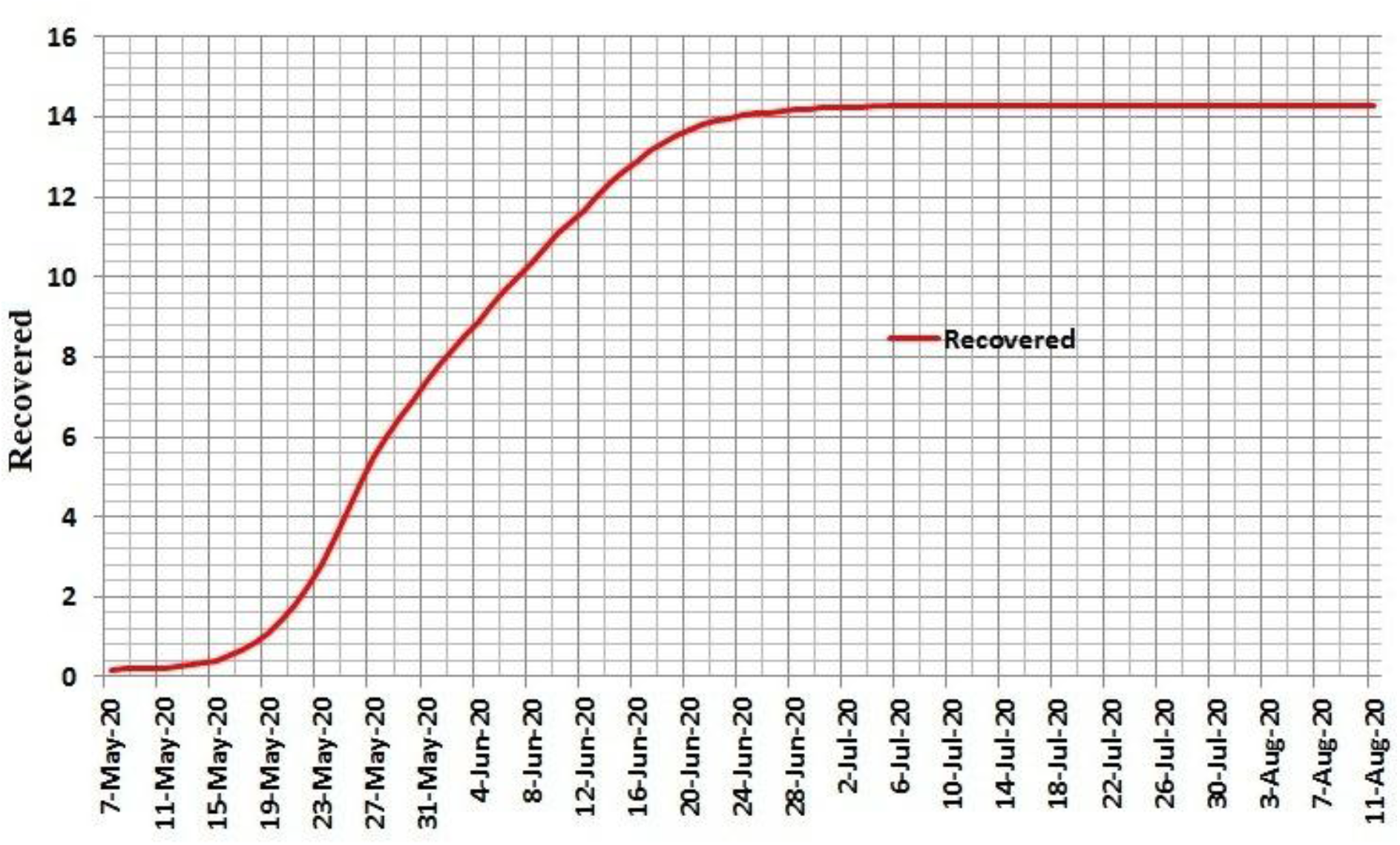
Recovered cases of COVI-2019 outbreak of India

The maximum number of infectives cases (*I*_max_) of COVID-2019 outbreak of India can be calculated using equation (17) is as follows:

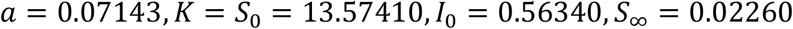

Then the ratio 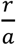 can be calculated using equation (15) i.e. 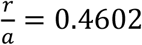. Therefore *r* = 0.4602 × 0.07143 = 0.32872086.

Hence *I*_max_ = 7.9835. Here, we have multiply by 100000 in *I*_max_ to get maximum number of infectives cases of COVID-2019 outbreak of India because 100000 is the normalization factor of this proposed study. Therefore *I*_max_ = 7.9835 × 10000 = 798350 which is the real data pointing at 782000 in table 1. From this table, we have seen that maximum number of infectives cases of CODID-2019 is 782000. This value is near by the *I*_max_. Therefore, we have seen that there will be a maximum outbreak of COVID-2019 in India on 25 May 2020, then it will decrease continuously till the first week of August 2020.

The reproductive number of COVID-2019 outbreak can be also calculated on initial, pick, end of COVID-2019 outbreak and any time during epidemic of COVID-2019 of India. Here, there are some reproductive number calculations are given below:

1. Initial level of COVID-2019: 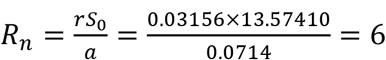
2. Pick level (maximum of COVID-2019): R_n_ = 13.57410 × 0.41602 = 6.2468

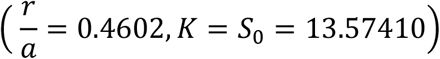
3. End level of COVID-2019: R_n_ = 0.0226 × 0.4602 = 0.001040

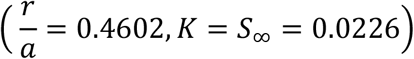

From above calculation, we have observed that the if reproductive number is greater than one then the COVID-2019 increasing continuously at pick/maximum level Case-1 and case-2) and if reproductive number is less than one then the COVID-2019 is died off (case-3). However, the reproduction number of COVID-2019 has been calculated by epidemiological scientists all over the world.

In the study presented, we get the following observation:

1. The outbreak of COVID-2019 will be at its peak on 25 May 2020 in India, after which the outbreak of this epidemic will continue to work slowly.
2. The outbreak of COVID-2019 in India will be till the first or second week of August 2020, after which the epidemic will end.
3. The initial stage of COVID-2019, the reproductive number of this outbreak of India is 6.

## 4. Conclusion

Based on the data as of 9 May 2020 in the study presented, the SIR model indicates that COVID-2019 outbreak will be at its peak in India by 25 May 2020 or by the end of May. On the basis of this study, we can say that after the end of May 2020, the outbreak of this epidemic will start working slowly and by the first week of August 2020, the outbreak of this epidemic will be towards end. On the basis of the data obtained by this model, it would be wrong to say that the COVID-2019 outbreak in India will go on because people here today are neither following social distancing nor applying their face masks. Hence this epidemic threat is very high risk in India. This study also shows that if locking, social distancing and masks etc. are used properly in India, then the outbreak of COVID-2019 epidemic can be almost eliminated in the first or second week of August 2020. This proposed study is very useful for the future prediction of outbreak of COVID-2019. This proposed SIR model will automatically estimates the number of cases of weekly, bi-weekly, month and even year. Therefore, we can say that the Indian government and doctors can maintain a check on hospital facilities, necessary supplies for new patients, medical aid and isolation for next week or in future.

## Data Availability

Dear Sir,
I have taken the COVID-outbreak data from Indian Council of Medical Research (ICMR) of India on 7 May 2020 and some data from WHO web sites on 15 May 2020. The websites are given below:

https://www.icmr.gov.in/

https://www.who.int/emergencies/diseases/novel-coronavirus-2019/situation-reports/

